# Trends in Prevalence of Cannabis Use Disorders among U.S. Veterans with and without Psychiatric Disorders Between 2005 and 2019

**DOI:** 10.1101/2023.05.24.23290358

**Authors:** Ofir Livne, Carol A. Malte, Mark Olfson, Melanie M. Wall, Katherine M. Keyes, Charles Maynard, Jaimie L. Gradus, Andrew J. Saxon, Silvia S. Martins, Salomeh Keyhani, Yoanna McDowell, David S. Fink, Zachary L. Mannes, Sarah Gutkind, Deborah S. Hasin

## Abstract

**Objective:** Cannabis use disorders (CUD) are increasing among U.S. adults and are more prevalent among cannabis users with comorbid psychiatric disorders. Changing cannabis laws, increasing cannabis availability, and higher potency cannabis may have recently placed cannabis users with psychiatric disorders at disproportionately increasing risk for CUD. The authors used Veterans Health Administration (VHA) data to examine whether trends in CUD prevalence among VHA patients differ by whether they have psychiatric disorders.

**Methods:** VHA electronic health records from 2005 to 2019 (n range=4,332,165-5,657,277) were used to identify overall and age-specific (<35, 35-64, ≥65 years) trends in prevalence of CUD diagnoses among patients with depressive, anxiety, PTSD, bipolar, or psychotic-spectrum disorders, and to compare these to corresponding trends among patients without any of these disorders. Given transitions in ICD coding, differences in trends were tested within two periods: 2005–2014 (ICD-9-CM) and 2016–2019 (ICD-10-CM).

**Results:** Greater increases in prevalence of CUD diagnoses were observed in veterans with, compared to without, psychiatric disorders (2005-2014: difference in prevalence change=1.91%, 95% CI=1.87%-1.96%; 2016-2019: 0.34%, 95% CI=0.29%-0.38%). Disproportionate increases in CUD prevalence among those with psychiatric disorders were greatest in veterans ages <35 between 2005-2014, and in those ages ≥65 between 2016-2019. Among patients with psychiatric disorders, greatest increases in CUD prevalences were observed in those with bipolar and psychotic-spectrum disorders.

**Conclusions:** Results highlight disproportionately increasing disparities in risk of CUD among VA patients with common psychiatric disorders. Greater public health and clinical efforts are needed to monitor, prevent and treat CUD among this population.

## INTRODUCTION

Many individuals use cannabis without evident harm. However, regular or heavy users are at increased risk of numerous cannabis-related health consequences^1-3^, including about 20-33% of users who develop cannabis use disorder (CUD)^4^. CUD is associated with impaired functioning, psychosocial impairments, and health problems^5,6^. Over the last two decades, non-medical use of cannabis has increased^7,8^, and the prevalence of adult CUD has increased in the U.S. general population^9,10^, in national samples of clinical populations^11,12^, and in U.S. veterans^13,14^. Identifying characteristics that are disproportionately increasing risk for medically recognized CUD over time is an important public health priority. Because CUD and psychiatric disorders are associated^6,15-17^, the presence of a psychiatric disorder may be one such characteristic.

In a survey of U.S. adults, nearly half believed that cannabis use is beneficial for stress, anxiety, or depression^18^, consistent with self-reported reasons for use^19^. However, reviews and meta-analyses^20,21^, suggest that using cannabinoids to treat psychiatric conditions increases the risk for adverse events while not providing significant benefits for depression, most anxiety conditions, posttraumatic stress disorder (PTSD) or psychotic disorders. Nevertheless, states that have legalized medical cannabis use generally permit its use for psychiatric conditions. Given the rapidly evolving U.S. cannabis landscape, including increasingly permissive attitudes and laws, increasing availability, and increasingly potent cannabis products^22,23^, people with psychiatric disorders may be a group whose risk for CUD is increasing disproportionately over time. If so, greater public health and clinical efforts should be focused on identifying, preventing and treating CUD among those with psychiatric disorders. Few sources of data are available on trends over time in CUD that also provide detailed information on diagnoses of specific psychiatric disorders.

The Veterans Health Administration (VHA) is the largest integrated healthcare system in the U.S. Within the VHA, a high proportion of patients are affected by psychiatric disorders^24^, and rates of CUD diagnoses have increased considerably since 2005^13,14^. The VHA electronic health record (EHR) database can be leveraged to compare trends in diagnoses of CUD between patients with and without comorbid psychiatric disorders. Using yearly VA EHR data from 2005-2019, we compared CUD trends among patients with and without common psychiatric disorders, including depressive, anxiety, posttraumatic stress, bipolar, and psychotic-spectrum disorders. Because of known age differences in the risk of CUD^6,17^, we also examined trends in CUD diagnoses across these psychiatric disorders by age group (<35, 35-64, ≥65).

## METHODS

### Sample & Procedure

Data from 1/1/2005 to 12/31/2019 were obtained through the Veterans Affairs (VA) Corporate Data Warehouse, a data repository for all care provided at VHA facilities or paid for by the VHA. Veterans ≥18 years with ≥1 VHA primary care, emergency department, or mental health visit in a given calendar were included in the study populations, except those in hospice/palliative care or residing outside the 50 states or Washington DC. Fourteen study populations were created, one for each year from 2005-2014 (n’s=4,332,165-5,383,712) and from 2016-2019 (n’s=5,508,670-5,657,277). (Due to the 2015 transition from ICD-9-CM to ICD-10-CM and methodological effects on rates across this transition, as described in detail elsewhere^14,25^, we excluded this year). Study approvals and waivers/exemptions of informed consent was granted by New York State Psychiatric Institute, VA Puget Sound and VA New York Harbor Healthcare Systems Institutional Review Boards.

### Measures

#### Cannabis use disorder diagnoses

The primary outcome was a provider-made CUD diagnosis given at ≥1 outpatient or inpatient encounter within a calendar year. We used ICD-9-CM from 2005-2014 (305.2X, abuse; 304.3X, dependence), and ICD-10-CM from 2016-2019 (F12.1X, abuse; F12.2X, dependence). We combined abuse and dependence because their criteria are unidimensional. As was done previously^14,25^, remission and unspecified CUD diagnoses were excluded.

#### Psychiatric disorders

ICD-9-CM or ICD-10-CM psychiatric disorder diagnoses indicated disorders in the following 5 categories: (1) depressive disorders; (2) anxiety disorders; (3) posttraumatic stress disorder (PTSD); (4) bipolar disorders; and (5) psychotic-spectrum disorders (for codes, see Table S1). All codes indicating disorders in remission were excluded. The 5 categories were selected because they include the most common psychiatric disorders recorded in the VHA EHR. For each category, a dichotomous variable was created indicating if patients were diagnosed with any of the disorders within that category in a given year, 2005-2014 (ICD-9-CM) and 2016-2019 (ICD-10-CM). These categories were not created to be mutually exclusive, so patients could be included in more than one category if they meet criteria for multiple conditions. These 5 dichotomous variables were used to create a binary summary variable indicating if patients were positive for ≥1 of the 5 categories within each year, hereafter termed “any psychiatric disorder”. To explore the sensitivity of the findings to comorbidity, we also created 5 analogous psychiatric disorder categories that were mutually exclusive, with each category including only the subset of patients who are diagnosed with a disorder in that given category and not diagnosed with disorders in any of the other 4 categories.

#### Covariates

Covariates included sex (male, female), age (<35 years, 35-64 years, ≥65 years), and race/ethnicity (non-Hispanic White, non-Hispanic Black, Hispanic; other/multiple races, and unknown).

### Statistical analysis

Following our previous work^14^, analyses were performed on the overall sample and then stratified by age (<35, 35-64, ≥65). Demographic characteristics, unadjusted prevalence of CUD and unadjusted prevalence of any psychiatric disorder and the 5 psychiatric disorder categories are presented for the overall sample and stratified by age group for study years 2005, 2014, 2016, and 2019. Due to the methodological issues created by the 2015 mid-year transition from ICD-9-CM to ICD-10-CM, we excluded this year and examined adjusted trends in CUD within two periods: 2005-2014 (ICD-9-CM) and 2016-2019 (ICD-10-CM). To test for changes in CUD over time by any psychiatric disorder, multivariable logistic regression models were used that included categorical study year, any psychiatric disorder (yes, no) and an interaction term for any psychiatric disorder and study year, adjusted for sex, race/ethnicity, and age in three categories as above. Separate multivariable logistic regression models were then run stratified by age group, adjusting for, sex, race/ethnicity, and continuous age. The predicted diagnostic prevalence of CUD diagnoses (i.e., adjusted proportions) in each year, the change in prevalence, the difference in those trends between those with and without any psychiatric disorder and the associated 95% confidence intervals (CI) were obtained from the margins command for the fitted logistic regression model. CIs were based on standard statistical methods that assume normal sampling distributions. While full-census data such as VA EHR data do not necessarily require uncertainty estimates, we present them to indicate the precision of our prevalence estimates.

Trends in CUD prevalence between 2005-2014 and 2016-2019 were then examined within each of the 5 diagnostic categories using logistic regression models including study year, adjusted for demographic covariates. Each model was limited to patients diagnosed with the specific condition to examine within-group trends. These diagnostic categories were not exclusive, such that patients were included in more than 1 category if they met criteria for multiple conditions. Then, to check robustness of the findings to potential influences of comorbidity, similar models were run for each of the 5 categories using the variables indicating patients diagnosed only with the specific category and no other categories. Stata v.17 was used for all analyses.

## RESULTS

### Psychiatric disorders and demographic characteristics

Prevalences (2005, 2019) of any psychiatric disorder and each of the 5 specific disorder categories are presented for the overall sample and by age categories in Table S2a. From 2005-2019, the prevalence of any psychiatric disorder increased from 19.0% to 29.2%. The most common diagnoses were depressive disorders (2005: 10.5%, 2019: 16.1%) and PTSD (2005: 6.2%, 2019: 14.8%). Overall and age-stratified demographic characteristics (2005, 2019) are presented for the entire sample, and for patients with and without psychiatric disorders in Table S2b. In the overall sample, patients were primarily male (2005: 95.0%, 2019: 90.8%) and non-Hispanic White (2005: 78.7%, 2019: 70.3%; Table S2b). About half were age ≥65 years (2005: 49.2%, 2019: 51.9%). In both time periods, patients with and without psychiatric disorders were primarily male (2005: 92.2%, 95.7%, respectively; 2019: 85.6%, 93.0%, respectively), and non-Hispanic White (2005: 75.9%, 79.3%, respectively; 2019: 65.2%, 72.5%, respectively).

### CUD

In the overall sample, the adjusted prevalence of ICD-9-CM CUD diagnoses increased two-fold from 0.85% in 2005 to 1.87% in 2014. As described and discussed in detail previously^14,25^, the ICD-9-CM to ICD-10-CM transition led to an artifactual decrease in CUD prevalence in 2016 due to provider coding practices and EHR procedures. Between 2016 and 2019, the adjusted prevalence of ICD-10-CM CUD increased from 1.62% to 1.92%.

### Overall trends in prevalence of CUD in veterans with and without any psychiatric disorder

#### 2005-2014 (Table 1; Figure 1)

In 2005, the prevalence of ICD-9-CM CUD was greater in patients with any psychiatric disorder than in other patients (2.48% and 0.35%, respectively. Over time, the adjusted overall prevalence of CUD diagnoses increased among patients with and without any psychiatric disorder. In patients with any psychiatric disorder, the prevalence of ICD-9-CM CUD diagnoses increased by 2.22%, reaching 4.70% in 2014. In patients without any psychiatric disorder, the prevalence of ICD-9-CM CUD diagnoses increased by 0.31%, reaching 0.66% in 2014. The increase in patients with any psychiatric disorder was significantly greater than in patients without these disorders (difference in prevalence change=1.91%; 95% CI=1.87-1.96).

**Table 1.** Overall and Age-Stratified Change in the Diagnostic Prevalence of CUD by Any Psychiatric Disorder^a^ in VA patients, 2005-2019

|  |  |  |  |  |  |  |
| --- | --- | --- | --- | --- | --- | --- |
|  | **Adjusted Prevalence<sup>b</sup>** |  |  |  |  |  |
|  | **ICD-9-CM CUD diagnoses** |  |  | **ICD-10-CM CUD diagnoses** |  |  |
|  | **2005**<br>**% (95% CI)** | **2014**<br>**% (95% CI)** | **Change**<br>**(95% CI)** | **2016**<br>**% (95% CI)** | **2019**<br>**% (95% CI)** | **Change**<br>**(95% CI)** |
| **Any Psychiatric Disorder** |  |  |  |  |  |  |
| **Overall** |  |  |  |  |  |  |
| No | 0.35 (0.34-0.35) | 0.66 (0.65-0.66) | 0.31 (0.30-0.32) | 0.51 (0.50-0.51) | 0.57 (0.56-0.58) | 0.06 (0.05-0.07) |
| Yes | 2.48 (2.45-2.51) | 4.70 (4.67-4.74) | 2.22 (2.18-2.27) | 4.28 (4.25-4.31) | 4.68 (4.65-4.71) | 0.40 (0.36-0.44) |
| Difference in prevalence | 2.13 (2.10-2.16) | 4.05 (4.02-4.08) | **1.91 (1.87-1.96)** | 3.77 (3.74-3.81) | 4.11 (4.08-4.14) | **0.34 (0.29-0.38)** |
| **Age Groups <35** |  |  |  |  |  |  |
| No | 0.57 (0.53-0.61) | 1.01(0.98-1.05) | 0.44 (0.39-0.50) | 0.89 (0.86-0.93) | 0.93 (0.89-0.96) | 0.04 (-0.01-0.08) |
| Yes | 5.56 (5.53-5.80) | 9.67 (9.54-9.81) | 4.11 (3.84-4.38) | 9.28 (9.15-9.41) | 9.75 (9.62-9.88) | 0.47 (0.29-0.65) |
| Difference in prevalence | 4.99 (4.76-5.23) | 8.66 (8.52-8.80) | **3.66 (3.39-3.94)** | 8.39 (8.25-8.52) | 8.82 (8.69-8.96) | **0.43 (0.24-0.62)** |
| **35-64** |  |  |  |  |  |  |
| No | 0.69 (0.67-0.70) | 1.13(1.11-1.15) | 0.44 (0.42-0.46) | 0.82 (0.80-0.83) | 0.81 (0.79-0.82) | -0.01 (-0.03-0.01) |
| Yes | 4.14 (4.08-4.19) | 6.99 (6.93-7.05) | 2.85 (2.78-2.93) | 6.04 (5.99-6.10) | 6.39 (6.34-6.44) | 0.35 (0.27-0.43) |
| Difference in prevalence | 3.45 (3.39-3.50) | 5.86 (5.80-5.92) | **2.41 (2.33-2.49)** | 5.23 (5.17-5.28) | 5.59 (5.53-5.64) | **0.36 (0.28-0.44)** |
| **≥65** |  |  |  |  |  |  |
| No | 0.02 (0.02-0.02) | 0.19 (0.18-0.19) | 0.17 (0.16-0.17) | 0.17 (0.17-0.18) | 0.30 (0.29-0.31) | 0.13 (0.12-0.14) |
| Yes | 0.16 (0.15-0.18) | 1.16 (1.14-1.18) | 0.99 (0.96-1.02) | 1.27 (1.24-1.29) | 2.04 (2.00-2.07) | 0.77 (0.73-0.81) |
| Difference in prevalence | 0.14 (0.13-0.16) | 0.97 (0.95-1.00) | **0.83 (0.80-0.86)** | 1.10 (1.07-1.12) | 1.74 (1.70-1.77) | **0.64 (0.60-0.68)** |
| <sup>a</sup> Dichotomous psychiatric summary variable indicating if patients were positive for any disorder from one or more of the 5 categories (depressive disorders, anxiety disorders, PTSD, bipolar disorders, psychotic-spectrum disorders) each year, 2005-2014 and 2016-2019.<br><sup>b</sup> Models include year, mental health condition, year X mental health condition, sex, race/ethnicity, continuous age.<br>Statistical significance indicated by positive difference in differences and confidence intervals that did not include 0.<br>Bolded values indicate statistically significant differences between those with psychiatric disorders versus those without psychiatric disorders. |  |  |  |  |  |  |

**Figure 1.**
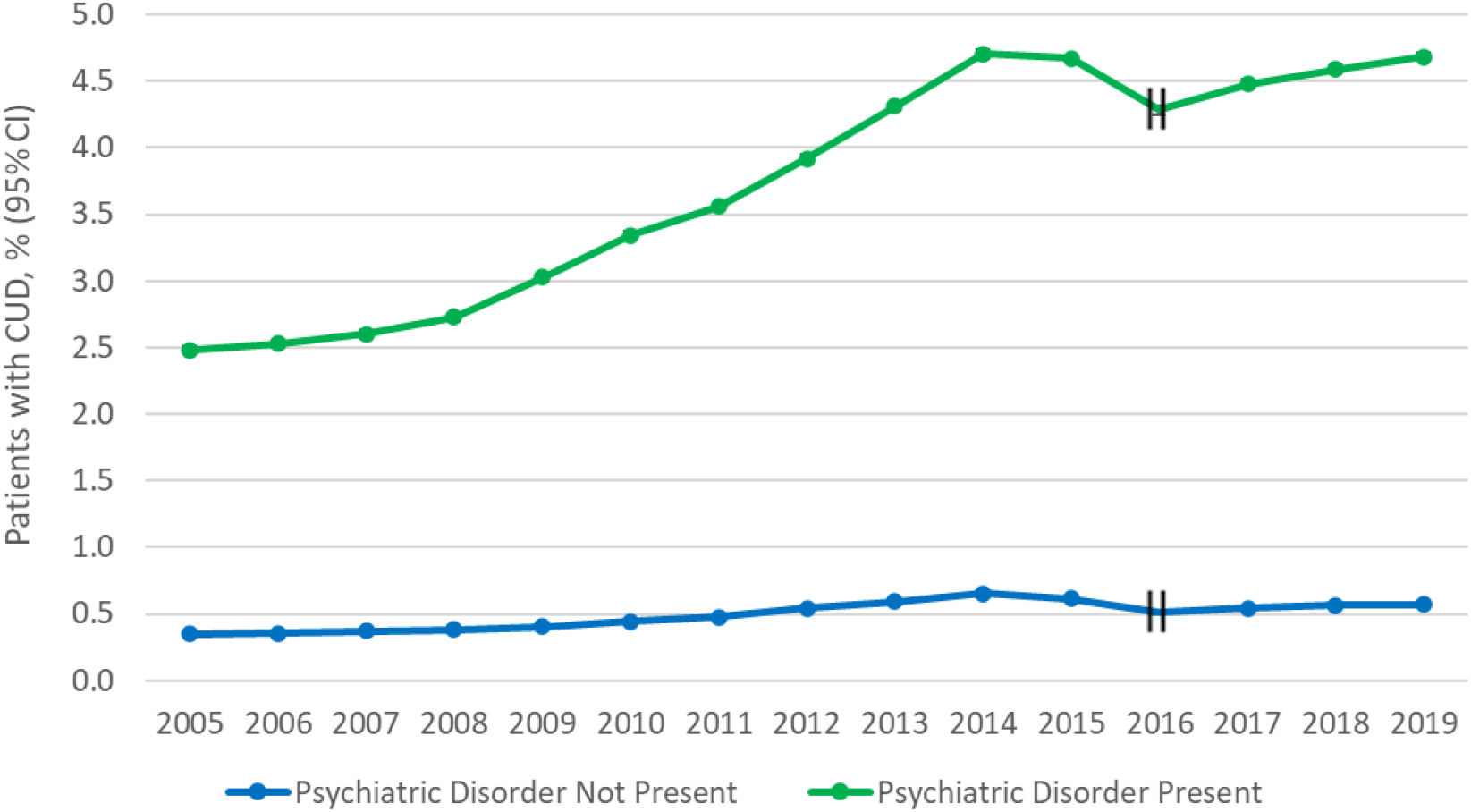
Overall Trends in Prevalence of CUD by Psychiatric Disorders (PSYCH)^a^ in VA Patients, 2005-2019. ^a^ Dichotomous psychiatric summary variable indicating if patients were positive for any disorder from ≥1 of the 5 categories (depressive disorders, anxiety disorders, PTSD, bipolar disorders, psychotic-spectrum disorders) each year, 2005-2014 and 2016-2019. Hash marks at 2015 indicate that this year was not included in models due to a change in ICD coding.

#### 2016-2019 (Table 1; Figure 1)

In 2016, the prevalence of ICD-10-CM CUD was greater in patients with any psychiatric disorder than in other patients (4.28% and 0.51%, respectively). Over time, the adjusted overall prevalence of ICD-10-CM CUD diagnoses increased among patients with and without any psychiatric disorder. In patients with any psychiatric disorder, the prevalence of ICD-10-CM CUD diagnoses increased by 0.40%, reaching 4.68% in 2019. In patients without any psychiatric disorder, the prevalence of ICD-10-CM CUD diagnoses increased by 0.06%, reaching 0.57% in 2019, a significantly greater absolute increase among patients with any psychiatric disorder of 0.34% (95% CI=0.29-0.38).

### Age-stratified trends in prevalence of CUD in veterans with and without any psychiatric disorder

#### 2005-2014 (Table 1; Figures S1A-S1C)

Across all age groups, patients with any psychiatric disorder had significantly greater increases in prevalence of CUD diagnoses than other patients. Between 2005-2014, the greatest absolute increases among patients with any psychiatric disorder occurred in adults <35 years (difference in prevalence change=3.66%, 95% CI=3.39-3.94), followed by those age 35-64 (difference in prevalence change =2.41%, 95% CI=2.33-2.49) and age ≥65 (difference in prevalence change =0.83%, 95% CI=0.80-0.86).

#### 2016-2019 (Table 1; Figures S1A-S1C)

Greatest differences in absolute increases in percent change in prevalence of CUD diagnoses between patients with and without any psychiatric disorder occurred in those age ≥65 years (difference in prevalence change =0.64%, 95% CI=0.60-0.68), followed by those age <35 years (difference in prevalence change=0.43%, 95% CI=0.24-0.62), and age 35-64 (difference in prevalence change =0.36%, 95% CI=0.28-0.44).

### Trends in prevalence of CUD in veterans with specific psychiatric disorders

#### 2005-2014

Within the 5 psychiatric disorder categories that were not mutually exclusive (comorbidity between the categories permitted), the prevalence of ICD-9-CM CUD ranged considerably (Table 2a; Figure 2a), with lowest prevalence among patients in the depressive disorder category (2005: 3.16%; 2014: 5.74%) and highest prevalences in the bipolar (2005: 6.65%; 2014: 12.31%) and psychotic-spectrum categories (2005: 5.07%; 2014: 11.08%). The increase in ICD-9-CM CUD diagnosis prevalence between 2005 and 2014 were significant within all 5 of the categories, with the smallest among those with depressive disorders (2.58%, 95% CI=2.51-2.65), followed by those with PTSD (2.93%, 95% CI=2.84-3.02), and those with anxiety disorders (3.49%, 95% CI=3.38-3.60). The increases in CUD prevalence were greater in those with bipolar or psychotic-spectrum disorder: 5.66% (95% CI=5.40-5.91) for bipolar disorders and 6.01% (95% CI=5.79-6.23) for psychotic-spectrum disorders.

**Table 2.**
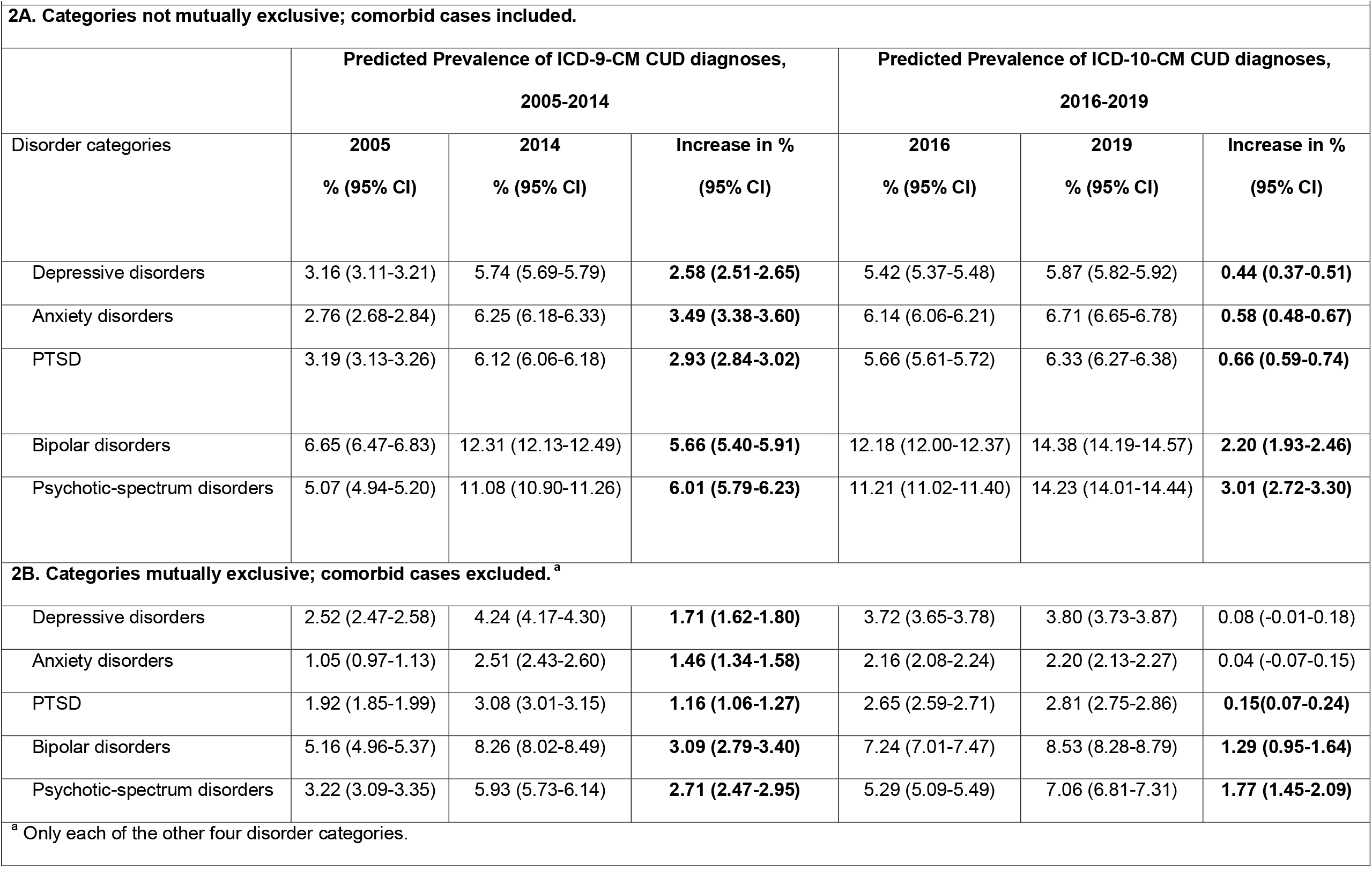
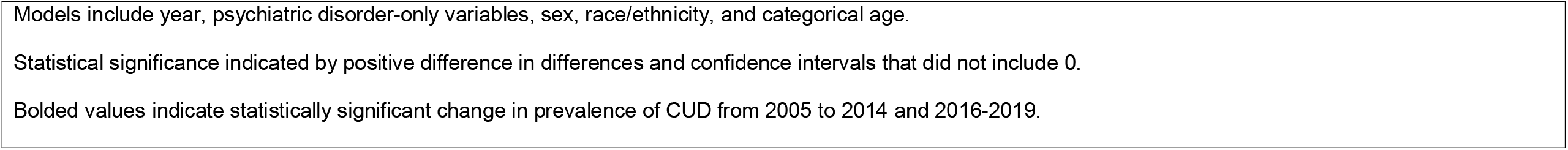
Change in the Predicted Diagnostic Prevalence of CUD by 5 Categories of Psychiatric Disorders in VA patients, 2005-2019

**Figure 2a.**
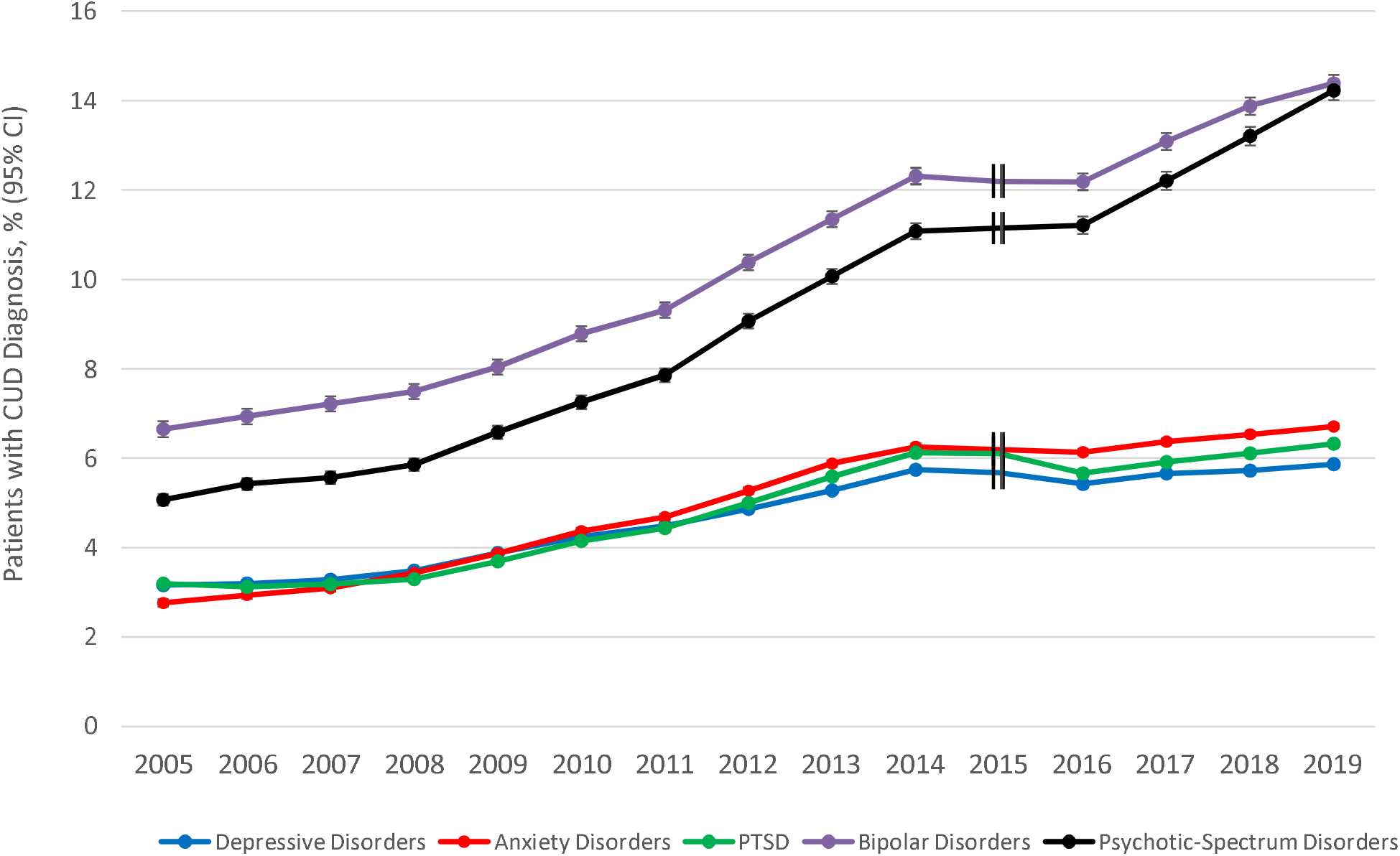
Overall Trends in Prevalence of CUD by 5 Categories of Psychiatric Disorders that are not mutually exclusive (comorbid cases included) in VA patients, 2005-2019 Hash marks at 2015 indicate that this year was not included in models due to a change in ICD coding.

Using the 5 psychiatric disorder categories that were mutually exclusive (indicating only the subset of patients in each category that had no comorbidity with the other four categories; Table 2b, Figure 2b), the overall prevalences of ICD-9-CM and ICD-10-CM CUD were lower than when the comorbid cases were included. However, CUD prevalence increased significantly among patients within each of the 5 specific non-comorbid disorder categories.

**Figure 2b.**
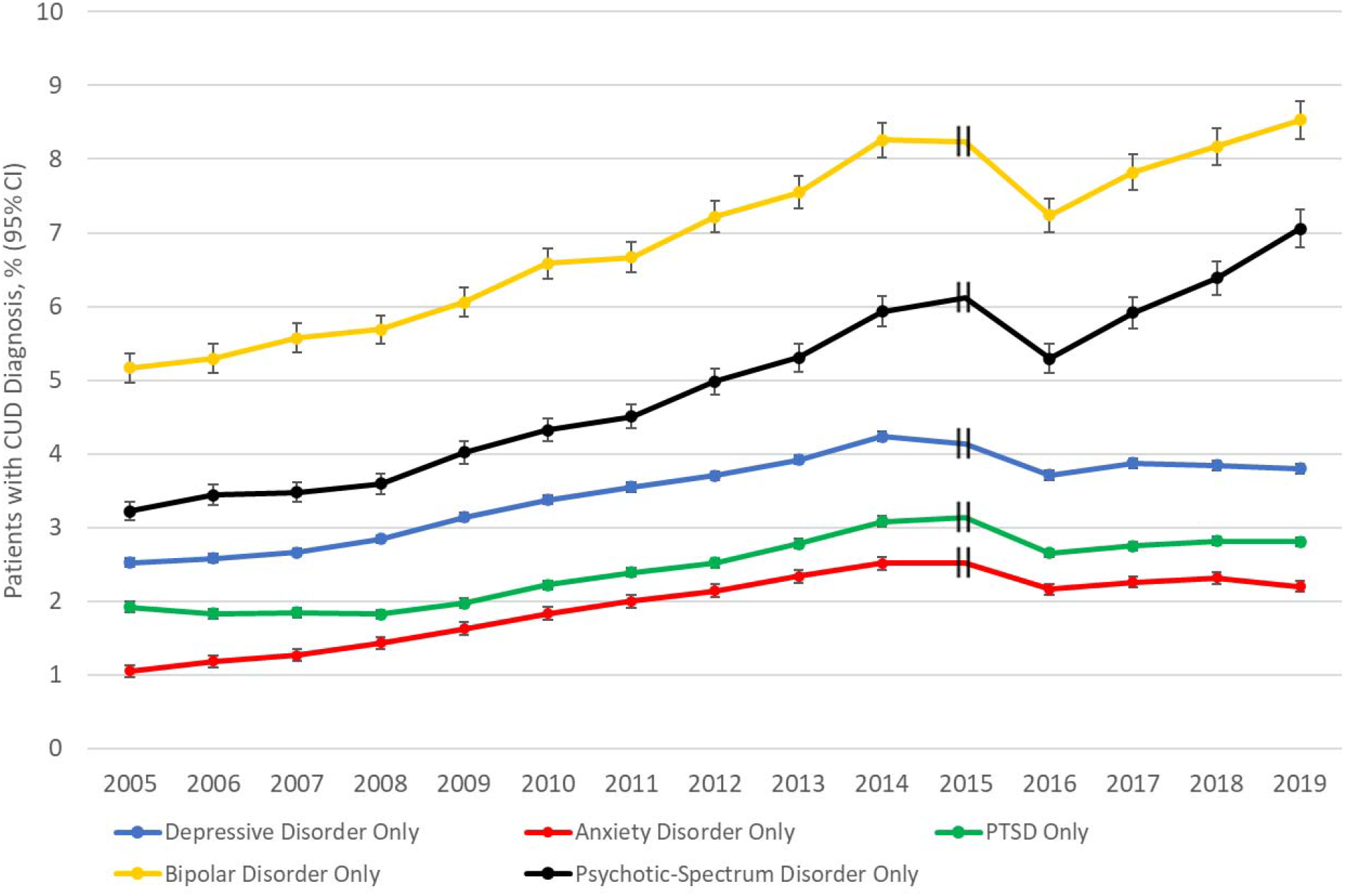
Overall Trends in Prevalence of CUD by 5 Categories of Psychiatric Disorders that are mutually exclusive (comorbid cases excluded) in VA patients, 2005-2019 Hash marks at 2015 indicate that this year was not included in models due to a change in ICD coding.

#### 2016-2019

Within the 5 psychiatric disorder categories defined as non-mutually exclusive (comorbidity between the categories permitted), the prevalence of ICD-10-CM CUD ranged considerably (Table 2a; Figure 2a), although the ranking CUD prevalence by disorder categories was similar to 2005-2014. The lowest prevalence of ICD-10-CM CUD was among patients in the depressive disorder category (2016: 5.42%; 2019: 5.87%). The highest were in the bipolar (2016: 12.18%; 2019: 14.38%) and psychotic-spectrum categories (2016:11.21%; 2019: 14.23%). Increases in ICD-10-CM CUD prevalence between 2016 and 2019 were smallest in patients with depressive disorders (0.44%, 95% CI=0.37-0.51), followed by anxiety disorders (0.58%, 95% CI=0.48-0.67) and PTSD (0.66%, 95% CI=0.59-0.74). Increases in CUD prevalence were greatest in patients with bipolar (2.20%, 95% CI=1.93-2.46) and psychotic-spectrum disorder (3.01%, 95% CI=2.72-3.30).

Using the 5 psychiatric disorder categories that were mutually exclusive, increases in ICD-10-CM CUD prevalence among patients with depressive or anxiety disorders were slight and not significantly different from zero (Table 2b; Figure 2b). However, the prevalence of ICD-10-CM CUD increased significantly among patients with PTSD, bipolar and psychotic-spectrum disorders.

### Age-stratified trends in prevalence of CUD in veterans by specific disorder categories

#### 2005-2014 (Table S3; Figures S2A-S2C)

CUD prevalence in patients with each of the disorder categories increased across age categories, with greatest increases observed in patients <35 years, followed by those age 35-64, and those age ≥65.

#### 2016-2019 (Table S3; Figures S2A-S2C)

CUD prevalence in patients with depressive disorders and anxiety disorders increased in those ages 35-64, and ages ≥65, but not in those ages <35. CUD prevalence in patients with PTSD, bipolar disorders, and psychotic-spectrum disorders increased in patients across all age groups.

## DISCUSSION

This study used VHA EHR data to examine overall and age-specific differences in time trends in the annual prevalence of CUD among VHA patients with and without psychiatric disorders over two periods (2005-2014, 2016-2019). CUD rates more than doubled among all VHA patients from 2005-2019, with disproportionately greater increases in patients with psychiatric disorders compared to those without. Among patients with psychiatric disorders, the greatest disparities occurred in those with bipolar and psychotic-spectrum disorders. Trends also varied by age groups – disparities in CUD rates between those with and without psychiatric disorders were greatest in veterans ages <35 years between 2005-2014 but greatest in those ages ≥65 between 2016-2019. Altogether, our findings show changes over time in the vulnerability of certain cannabis-using populations to CUD, i.e., those with psychiatric disorders, and identify age groups at higher risk that need to be closely monitored for CUD.

The increasing prevalence of CUD in the overall VHA patient population reported in this study generally support previous analyses of VHA^14^ samples, and some nationally representative U.S. survey reports^9,26-28^. Our findings that increasing prevalence of CUD was disproportionately greater among veterans with psychiatric disorders compared to other patients contributes novel findings to a small literature with limitations including analysis of cannabis use without CUD^29-32^; a focus mainly on mood, anxiety^29-31^ and psychotic disorders^32^, but not PTSD or bipolar disorders; and that rarely report estimated changes in prevalence of CUD among individuals with psychiatric conditions.

When comorbidity between the examined psychiatric disorder categories was permitted, the prevalence of CUD diagnoses increased disproportionately among patients diagnosed with each of the 5 psychiatric disorder categories during both study time periods compared to patients without psychiatric disorders. When examining only non-comorbid cases, results showed similar trends, albeit with a few exceptions. Notably, the disproportionate increases in the prevalence of CUD diagnoses were particularly striking among individuals with what may be the most severe conditions, bipolar and psychotic-spectrum disorders. Our results on disproportionate increases in comorbidity of CUD and psychotic-spectrum disorders are consistent with previous findings from a study of self-reported psychotic diagnoses in the general population^32^. However, our finding on patients with bipolar disorders is, to our knowledge, the first reported time trend for comorbidity of CUD with these disorders. With greater recreational legalization and increased availability, patients with bipolar or psychotic-spectrum disorders may be using cannabis in an attempt to self-medicate, even though evidence suggests that this is inadvisable^20,21^.

From 2005-2014, differences in CUD rates between those with and without psychiatric disorders were greatest in veterans ages <35 years. However, from 2016-2019, these trends shifted, with greatest differences in CUD rates observed in veterans ages ≥65. This may be partly driven by the dramatic rise in recent years in cannabis use among U.S adults ≥65, compared to younger age groups^33,34^. Additionally, many Vietnam-era veterans, now older adults, continue to experience high rates of psychiatric disorders including PTSD^35^, which is now legally authorized to treat with cannabis in many U.S. states and for which self-medication with cannabis is common^36^. Future studies need to examine whether changing state cannabis laws are differentially influencing patients with and without psychiatric disorders overall, or those with specific psychiatric diagnoses.

Limitations are noted. First, VA patients are largely White middle-aged males with high rates of medical disorders^37^. Our findings may not be generalizable to non-VA veterans, to women, or to the general population, although our trend results are largely consistent with other general population findings^29-31^. Second, as with other studies using EHR data, diagnoses were based on ICD patient encounter codes entered by VA providers rather than on research assessments. Therefore, diagnoses likely include some degree of provider error, though they likely disproportionately capture patients with more severe or conspicuous conditions. Third, the transition from ICD-9-CM to ICD-10-CM necessitated examining trends within two time periods: 2005-2014 and 2016-2019. Recording practices for some conditions are likely to have affected the prevalence estimates of CUD^14,25^ and some psychiatric disorders as well. Fourth, some psychiatric disorders may be of interest (e.g., attention deficit hyperactivity disorder), but were outside the scope of the present study. Finally, the VA EHR does not include measures of cannabis use such as frequency, route of administration or motive for use (recreational vs. medical); therefore, we cannot assess if these factors affected the observed trends.

## CONCLUSIONS

During a period of increasing cannabis use and CUD in the U.S., the prevalence of CUD was disproportionally increased among VHA patients with psychiatric disorders, especially among those with more severe psychiatric disorders. Despite evidence of potential harms from cannabis use, U.S. adults have become increasingly likely to perceive cannabis use as harmless^38,39^ and useful for treating conditions such as stress, anxiety or depression^18^. However, individuals with comorbid CUD and psychiatric disorders are at increased risk of functional impairments, and other harms^4-6^. Consequently, greater public health and clinical efforts are needed to systematically monitor risky cannabis use and CUD and to develop preventive and harm reduction strategies in these cannabis-using populations.

## Data Availability

The data in the present study are not publicly available.

## Acknowledgements

Supported by NIDA grant R01DA048860, the New York State Psychiatric Institute, and the VA Centers of Excellence in Substance Addiction Treatment and Education.

