## Supplemental Material for "Trends in Prevalence of Cannabis Use Disorders among U.S. Veterans with and without Psychiatric Disorders Between 2005 and 2019"

| **Table S1.** Psychiatric diagnoses and corresponding ICD-9, ICD-10 diagnostic codes. | | |
| --- | --- | --- |
| **Psychiatric Disorders** | **ICD-9 Code** | **ICD-10 Code** |
| Depressive Disorders | 296.20-296.25, 296.30-296.35, 296.82, 296.90, 300.4, 301.12, 311. | F32.0-F32.4, F32.8X, F32.9, F32.A, F33.0-F33.3, F33.41, F33.8, F33.9, F34.1, F34.8X, F34.9, F39. |
| Anxiety disorders | 300.0X, 300.2X | F40.X, F41.X |
| Posttraumatic Stress Disorder | 309.81 | F43.10, F43.12 |
| Bipolar Disorder | 296.00-296.05, 296.10-296.15, 296.40-296.45, 296.50-296.55, 296.60-296.65, 296.7, 296.80, 296.81, 296.89, 301.13 | F30.1X-F30.3, F30.8, F30.9, F34.0 |
| Psychotic Spectrum Disorder | 295.00-295.04, 295.10-295.14, 295.20-295.24, 295.30-295.34, 295.40-295.44, 295.60-295.64, 295.70-295.74, 295.80-295.84, 295.90-295.94, 297.X, 298.X | F20.0-F20.5, F20.81, F20.9, F21.-F25.X, F28., F29. |

| **Table S2a.** Adjusted Prevalences of Psychiatric Disorders in VA Patients, 2005 and 2019, overall and by age group. | | | | |
| --- | --- | --- | --- | --- |
|  | **2005** | | **2019** | |
|  | n | (%) | n | (%) |
| **All ages 18 years and older** | **N=4,332,165** | | **N=5,657,277** | |
| Any psychiatric disorder^a^ | 824,019 | (19.02) | 1,652,604 | (29.21) |
| Depressive disorders | 452,950 | (10.46) | 912,804 | (16.14) |
| Anxiety disorders | 162,551 | (3.75) | 574,621 | (10.16) |
| PTSD | 267,671 | (6.18) | 839,439 | (14.84) |
| Bipolar disorders | 72,681 | (1.68) | 128,146 | (2.27) |
| Psychotic-spectrum disorders | 117,671 | (2.72) | 99,132 | (1.75) |
| **<35 years** | **n=187,692** | | **n=482,749** | |
| Any psychiatric disorder^a^ | 37,912 | (20.20) | 203,386 | (42.13) |
| Depressive disorders | 20,571 | (10.96) | 108,160 | (22.41) |
| Anxiety disorders | 8,367 | (4.46) | 87,510 | (18.13) |
| PTSD | 12,817 | (6.83) | 108,788 | (22.54) |
| Bipolar disorders | 4,957 | (2.64) | 16,278 | (3.37 |
| Psychotic-spectrum disorders | 3,614 | (1.93) | 8,712 | (1.80 |
| **35-64 years** | **n=2,015,250** | | **n=2,235,902** | |
| Any psychiatric disorder | 566,190 | (28.10) | 838,489 | (37.50) |
| Depressive disorders | 305,977 | (15.18) | 472,542 | (21.13) |
| Anxiety disorders | 100,288 | (4.98) | 307,313 | (13.74) |
| PTSD | 218,010 | (10.82) | 427,606 | (19.12) |
| Bipolar disorders | 57,133 | (2.84) | 80,187 | (3.59) |
| Psychotic-spectrum disorders | 77,764 | (3.86) | 53,249 | (2.38) |
| **≥65 years** | **n=2,129,223** | | **n=2,938,626** | |
| Any psychiatric disorder^a^ | 219,917 | (10.33) | 610,729 | (20.78) |
| Depressive disorders | 126,402 | (5.94) | 332,102 | (11.30) |
| Anxiety disorders | 53,896 | (2.53) | 179,798 | (6.12) |
| PTSD | 36,844 | (1.73) | 303,045 | (10.31) |
| Bipolar disorders | 10,591 | (0.50) | 31,681 | (1.08) |
| Psychotic-spectrum disorders | 36,293 | (1.70) | 37,171 | (1.26) |
| ^a^ Positive for any disorder from ≥1 of the 5 categories: depressive disorders, anxiety disorders, PTSD, bipolar disorders, psychotic-spectrum disorders | | | | |

| **Table S2b.** VA Patient Demographics, overall and by presence or absence of one or more of the psychiatric disorders from the 5 most common categories, 2005 and 2019. | | | | | | | | | | | | |
| --- | --- | --- | --- | --- | --- | --- | --- | --- | --- | --- | --- | --- |
|  | **2005 (N=4,332,165)** | | | | | | **2019 (N=5,657,277)** | | | | | |
|  | Overall | | No Psychiatric Disorder | | Any Psychiatric Disorder^a^ | | Overall | | No Psychiatric Disorder | | Any Psychiatric Disorder^a^ | |
|  | n | (%) | n | (%) | n | (%) | n | (%) | n | (%) | n | (%) |
| **Overall** |  | | **n=3,508,146** | | **n=824,019** | |  | | **n=4,004,673** | | **n=1,652,604** | |
| Sex |  |  |  |  |  |  |  |  |  |  |  |  |
| Female | 216,961 | (5.0) | 152,505 | (4.4) | 64,456 | (7.8) | 522,707 | (9.2) | 284,660 | (7.1) | 238,047 | (14.4) |
| Male | 4,115,199 | (95.0) | 3,355,636 | (95.7) | 759,563 | (92.2) | 5,134,565 | (90.8) | 3,720,009 | (93.0) | 1,414,556 | (85.6) |
| Race/Ethnicity |  |  |  |  |  |  |  |  |  |  |  |  |
| Non-Hispanic  White | 3,407,953 | (78.7) | 2,782,872 | (79.3) | 625,081 | (75.9) | 3,978,984 | (70.3) | 2,901,865 | (72.5) | 1,077,119 | (65.2) |
| Non-Hispanic  Black | 605,523 | (14.0) | 473,295 | (13.5) | 132,228 | (16.1) | 1,017,033 | (18.0) | 664,225 | (16.6) | 352,808 | (21.4) |
| Hispanic/Latino | 140,871 | (3.3) | 107,547 | (3.1) | 33,324 | (4.0) | 342,413 | (6.1) | 213,745 | (5.3) | 128,668 | (7.8) |
| Other/Multiple | 99,714 | (2.3) | 77,855 | (2.2) | 21,859 | (2.7) | 187,997 | (3.3) | 125,545 | (3.1) | 62,452 | (3.8) |
| Unknown | 78,104 | (1.8) | 66,577 | (1.9) | 11,527 | (1.4) | 130,850 | (2.3) | 99,293 | (2.5) | 31,557 | (1.9) |
| Age |  |  |  |  |  |  |  |  |  |  |  |  |
| <35 | 187,692 | (4.3) | 149,780 | (4.3) | 37,912 | (4.6) | 482,749 | (8.5) | 279,363 | (7.0) | 203,386 | (12.3) |
| 35-64 | 2,015,250 | (46.5) | 1,449,060 | (41.3) | 566,190 | (68.7) | 2,235,902 | (39.5) | 1,397,413 | (34.9) | 838,489 | (50.7) |
| 65+ | 2,129,223 | (49.2) | 1,909,306 | (54.4) | 219,917 | (26.7) | 2,938,626 | (51.9) | 2,327,897 | (58.1) | 610,729 | (37.0) |
| Age, M(SD) |  |  | 64.8 | (14.7) | 58.4 | (13.5) |  |  | 64.4 | (16.4) | 56.0 | (16.0) |
| **<35** |  | | **n=149,780** | | **n=37,912** | |  | | **n=279,363** | | **n=203,386** | |
| Sex |  |  |  |  |  |  |  |  |  |  |  |  |
| Female | 43,221 | (23.0) | 32,967 | (22.0) | 10,254 | (27.1) | 95,330 | (19.8) | 48,122 | (17.2) | 47,208 | (23.2) |
| Male | 144,471 | (77.0) | 116,813 | (78.0) | 27,658 | (73.0) | 387,419 | (80.3) | 231,241 | (82.8) | 156,178 | (76.8) |
| Race/Ethnicity |  |  |  |  |  |  |  |  |  |  |  |  |
| White | 110,641 | (59.0) | 85,123 | (56.8) | 25,518 | (67.0) | 286,132 | (59.3) | 163,776 | (58.6) | 122,356 | (60.2) |
| Black | 37,011 | (19.7) | 30,617 | (20.4) | 6,394 | (16.9) | 80,380 | (16.7) | 44,342 | (15.9) | 36,038 | (17.7) |
| Hispanic/Latino | 16,556 | (8.8) | 13,664 | (9.1) | 2,892 | (7.6) | 64,013 | (13.3) | 37,126 | (13.3) | 26,887 | (13.2) |
| Other/Multiple | 7,119 | (3.8) | 5,689 | (3.8) | 1,430 | (3.8) | 27,335 | (5.7) | 16,031 | (5.7) | 11,304 | (5.6) |
| Unknown | 16,365 | (8.7) | 14,687 | (9.8) | 1,678 | (4.4) | 24,889 | (5.2) | 18,088 | (6.5) | 6,801 | (3.3) |
| Age, M(SD) |  |  | 28.0 | (3.9) | 28.7 | (3.8) |  |  | 29.5 | (3.3) | 30.2 | (3.1) |
| **35-64** |  | | **n=1,449,060** | | **n=566,190** | |  | | **n=1,397,413** | | **n=838,489** | |
| Sex |  |  |  |  |  |  |  |  |  |  |  |  |
| Female | 86,773 | (6.0) | 86,773 | (6.0) | 48,041 | (8.5) | 181,474 | (13.0) | 181,474 | (13.0) | 164,740 | (19.7) |
| Male | 1,362,285 | (94.0) | 1,362,285 | (94.0) | 518,149 | (91.5) | 1,215,937 | (87.0) | 1,215,937 | (87.0) | 673,748 | (80.4) |
| Race/Ethnicity |  |  |  |  |  |  |  |  |  |  |  |  |
| White | 1,000,537 | (69.1) | 1,000,537 | (69.1) | 406,905 | (71.9) | 816,049 | (58.4) | 816,049 | (58.4) | 489,828 | (58.4) |
| Black | 305,575 | (21.1) | 305,575 | (21.1) | 109,189 | (19.3) | 355,695 | (25.5) | 355,695 | (25.5) | 219,774 | (26.2) |
| Hispanic/Latino | 57,053 | (3.9) | 57,053 | (3.9) | 24,835 | (4.4) | 98,268 | (7.0) | 98,268 | (7.0) | 71,762 | (8.6) |
| Other/Multiple | 38,816 | (2.7) | 38,816 | (2.7) | 16,118 | (2.9) | 54,629 | (3.9) | 54,629 | (3.9) | 34,277 | (4.1) |
| Unknown | 47,079 | (3.3) | 47,079 | (3.3) | 9,143 | (1.6) | 72,772 | (5.2) | 72,772 | (5.2) | 22,848 | (2.7) |
| Age, M(SD) | 54.0 | (7.4) | 54.0 | (7.4) | 53.6 | (6.9) | 52.4 | (8.6) | 52.4 | (8.6) | 50.2 | (9.0) |
| **≥65** |  | | **n=1,909,306** | | **n=219,917** | |  | | **n=2,327,897** | | **n=610,729** | |
| Sex |  |  |  |  |  |  |  |  |  |  |  |  |
| Female | 32,765 | (1.7) | 32,765 | (1.7) | 6,161 | (2.8) | 55,064 | (2.4) | 55,064 | (2.4) | 26,099 | (4.3) |
| Male | 1,876,538 | (98.3) | 1,876,538 | (98.3) | 213,756 | (97.2) | 2,272,831 | (97.6) | 2,272,831 | (97.6) | 584,630 | (95.7) |
| Race/Ethnicity |  |  |  |  |  |  |  |  |  |  |  |  |
| White | 1,697,212 | (88.9) | 1,697,212 | (88.9) | 192,658 | (87.6) | 1,922,040 | (82.6) | 1,922,040 | (82.6) | 464,935 | (76.1) |
| Black | 137,103 | (7.2) | 137,103 | (7.2) | 16,645 | (7.6) | 264,188 | (11.4) | 264,188 | (11.4) | 96,996 | (15.9) |
| Hispanic/Latino | 36,830 | (1.9) | 36,830 | (1.9) | 5,597 | (2.6) | 78,351 | (3.4) | 78,351 | (3.4) | 30,019 | (4.9) |
| Other/Multiple | 33,350 | (1.8) | 33,350 | (1.8) | 4,311 | (2.0) | 54,885 | (2.4) | 54,885 | (2.4) | 16,871 | (2.8) |
| Unknown | 4,811 | (0.3) | 4,811 | (0.3) | 706 | (0.3) | 8,433 | (0.4) | 8,433 | (0.4) | 1,908 | (0.3) |
| Age, M(SD) | 75.8 | (6.4) | 75.8 | (6.4) | 75.8 | (6.6) | 75.7 | (7.5) | 75.7 | (7.5) | 72.6 | (5.8) |
| ^a^ Positive for any disorder from ≥1 of the 5 categories: depressive disorders, anxiety disorders, PTSD, bipolar disorders, psychotic-spectrum disorders | | | | | | | | | | | | |

| **Table S3.** Age-Stratified Change in the Diagnostic Prevalence of CUD by 5 Categories of Psychiatric Disorders^a^ in VA patients, 2005-2019. | | | | | | |
| --- | --- | --- | --- | --- | --- | --- |
|  | **Predicted Prevalence of ICD-9-CM CUD diagnoses,**  **2005-2014** | | | **Predicted Prevalence of ICD-10-CM CUD diagnoses,**  **2016-2019** | | |
|  | **2005**  **% (95% CI)** | **2014**  **% (95% CI)** | **Change (95% CI)** | **2016**  **% (95% CI)** | **2019**  **% (95% CI)** | **Change (95% CI)** |
| **Depressive disorders** | | | | | | |
| Age group | |  |  |  |  |  |
| <35 | 5.27 (4.96-5.58) | 10.34 (10.14-10.53) | **5.06 (4.70-5.43)** | 10.30 (10.10-10.49) | 10.14 (9.96-10.32) | -0.16 (-0.42-0.11) |
| 35-64 | 4.25 (4.18-4.32) | 6.92 (6.85-7.00) | **2.67 (2.57-2.78)** | 6.21 (6.13-6.28) | 6.58 (6.51-6.65) | **0.37 (0.27-0.48)** |
| ≥65 | 0.22 (0.19-0.26) | 1.72 (1.67-1.76) | **1.49 (1.44-1.55)** | 1.98 (1.93-2.03) | 2.99 (2.93-3.05) | **1.01 (0.93-1.09)** |
| **Anxiety disorders** | | | | | | |
| Age group | |  |  |  |  |  |
| <35 | 5.15 (4.68-5.63) | 10.61 (10.38-10.84) | **5.45 (4.93-5.98)** | 10.82 (10.59-11.05) | 10.74 (10.53-10.94) | -0.09 (-0.40-0.22) |
| 35-64 | 3.71 (3.59-3.82) | 7.47 (7.36-7.58) | **3.76 (3.60-3.92)** | 7.01 (6.90-7.12) | 7.56 (7.47-7.66) | **0.55 (0.41-0.70)** |
| ≥65 | 0.14 (0.10-0.18) | 1.64 (1.58-1.70) | **1.50 (1.43-1.58)** | 1.96 (1.89-2.04) | 3.14 (3.06-3.23) | **1.18 (1.07-1.29)** |
| **PTSD** | | | | | | |
| Age group | |  |  |  |  |  |
| <35 | 4.84 (4.47-5.20) | 10.41 (10.23-10.58) | **5.57 (5.16-5.97)** | 10.33 (10.15-10.50) | 11.69 (11.49-11.89) | **1.36 (1.10-1.63)** |
| 35-64 | 3.98 (3.90-4.07) | 6.82 (6.73-6.92) | **2.84 (2.71-2.96)** | 6.05 (5.97-6.14) | 6.61 (6.54-6.69) | **0.56 (0.45-0.67)** |
| ≥65 | 0.38 (0.29-0.47) | 2.08 (2.03-2.14) | **1.70 (1.60-1.81)** | 2.22 (2.16-2.27) | 3.77 (3.69-3.86) | **1.56 (1.46-1.65)** |
| **Bipolar disorders** | | | | | | |
| Age group | |  |  |  |  |  |
| <35 | 10.67 (9.82-11.53) | 22.09 (21.43-22.75) | **11.41 (10.33-12.49)** | 22.28 (21.62-22.93) | 23.78 (23.13-24.43) | **1.50 (0.58-2.43)** |
| 35-64 | 7.34 (7.13-7.55) | 13.10 (12.87-13.33) | **5.76 (5.45-6.07)** | 12.61 (12.37-12.85) | 14.77 (14.52-15.01) | **2.16 (1.82-2.50)** |
| ≥65 | 0.56 (0.39-0.72) | 3.74 (3.52-3.96) | **3.19 (2.91-3.46)** | 4.42 (4.18-4.67) | 6.71 (6.43-6.99) | **2.29 (1.92-2.66)** |
| **Psychotic disorders** | | | | | | |
| Age group | |  |  |  |  |  |
| <35 | 12.39 (11.32-13.46) | 30.24 (29.28-31.21) | **17.85 (16.41-19.29)** | 31.11 (30.13-32.09) | 33.66 (32.67-34.65) | **2.54 (1.15-3.94)** |
| 35-64 | 6.07 (5.91-6.23) | 12.89 (12.65-13.14) | **6.82 (6.53-7.12)** | 12.61 (12.34-12.87) | 15.69 (15.39-16.00) | **3.09 (2.68-3.49)** |
| ≥65 | 0.35 (0.27-0.44) | 2.82 (2.67-2.97) | **2.47 (2.29-2.64)** | 3.11 (2.94-3.28) | 4.85 (4.65-5.04) | **1.74 (1.48-2.00)** |
| ^a^ Categories are not mutually exclusive.  Models include year, mental health condition, year X mental health condition, sex, race/ethnicity, continuous age.  Statistical significance indicated by positive difference in differences and confidence intervals that did not include 0.  Bolded values indicate statistically significant change in prevalence of CUD from 2005 to 2014 and 2016-2019. | | | | | | |

**Figures S1a-S1c.** Age-Stratified Trends in Prevalence of CUD by Psychiatric Disorders (PSYCH)^1^ in VA Patients, 2005-2019

Figure S1a. Prevalence of CUD by Psychiatric Disorder, <35 Years

Patients with CUD, % (95% CI)

|  | Psychiatric Disorder Not Present | Psychiatric Disorder Present |
| --- | --- | --- |
| 2005 | 0.56878 | 5.56352 |
| 2006 | 0.56025 | 5.5693 |
| 2007 | 0.54861 | 5.43216 |
| 2008 | 0.54623 | 5.68455 |
| 2009 | 0.65623 | 6.56295 |
| 2010 | 0.73668 | 7.48796 |
| 2011 | 0.77858 | 7.90334 |
| 2012 | 0.88308 | 8.56861 |
| 2013 | 0.95246 | 9.18798 |
| 2014 | 1.01311 | 9.67192 |
| 2015 | 0.9866 | 9.55085 |
| 2016 | 0.8931 | 9.28151 |
| 2017 | 0.93741 | 9.64746 |
| 2018 | 0.95676 | 9.76139 |
| 2019 | 0.92918 | 9.75048 |

Figure S1b. Prevalence of CUD by Psychiatric Disorder, 35-64 Years

Patients with CUD, % (95% CI)

|  | Psychiatric Disorder Not Present | Psychiatric Disorder Present |
| --- | --- | --- |
| 2005 | 0.6874 | 4.13546 |
| 2006 | 0.71114 | 4.21695 |
| 2007 | 0.73731 | 4.33308 |
| 2008 | 0.75926 | 4.53119 |
| 2009 | 0.78759 | 4.96093 |
| 2010 | 0.8498 | 5.38697 |
| 2011 | 0.90221 | 5.6678 |
| 2012 | 0.99612 | 6.11371 |
| 2013 | 1.06411 | 6.54299 |
| 2014 | 1.13018 | 6.98981 |
| 2015 | 1.02214 | 6.79257 |
| 2016 | 0.81818 | 6.04369 |
| 2017 | 0.83773 | 6.25277 |
| 2018 | 0.83456 | 6.32648 |
| 2019 | 0.80618 | 6.39247 |

Figure S1c. Prevalence of CUD by Psychiatric Disorder, 65+ Years

Patients with CUD, % (95% CI)

|  | Psychiatric Disorder Not Present | Psychiatric Disorder Present |
| --- | --- | --- |
| 2005 | 0.02034 | 0.16496 |
| 2006 | 0.02199 | 0.20687 |
| 2007 | 0.03096 | 0.27237 |
| 2008 | 0.03999 | 0.32982 |
| 2009 | 0.04938 | 0.41825 |
| 2010 | 0.06669 | 0.52821 |
| 2011 | 0.08021 | 0.62047 |
| 2012 | 0.11158 | 0.75125 |
| 2013 | 0.1429 | 0.94564 |
| 2014 | 0.18535 | 1.15856 |
| 2015 | 0.19337 | 1.28888 |
| 2016 | 0.17132 | 1.26772 |
| 2017 | 0.21318 | 1.49999 |
| 2018 | 0.26152 | 1.76939 |
| 2019 | 0.29933 | 2.03639 |

^1^Dichotomous psychiatric summary variable indicating if patients were positive for any disorder from ≥1 of the 5 categories (depressive disorders, anxiety disorders, PTSD, bipolar disorders, psychotic-spectrum disorders*)* each year, 2005-2014 and 2016-2019.

Hatch marks at 2015 indicate that this year was not included in models due to a change in ICD coding

**Figures S2A-S2C.** Age-Stratified Trends in Prevalence of CUD by 5 Categories of Psychiatric Disorders^1^ in VA patients, 2005-2019.

Figure S2a. Trend in CUD by Psychiatric Disorders, <35 Years

Patients with CUD Diagnosis, % (95% CI)

|  | Depressive Disorders | Anxiety Disorders | PTSD | Bipolar Disorders | Psychotic-Spectrum Disorders |
| --- | --- | --- | --- | --- | --- |
| 2005 | 5.2724 | 5.15379 | 4.83664 | 10.6746 | 12.3907 |
| 2006 | 5.32141 | 5.5998 | 4.94125 | 11.8157 | 13.3549 |
| 2007 | 5.47607 | 5.74279 | 4.92804 | 12.1731 | 13.9011 |
| 2008 | 5.91233 | 6.27422 | 5.40191 | 12.3291 | 15.881 |
| 2009 | 7.03558 | 7.13108 | 6.46652 | 13.6732 | 17.7269 |
| 2010 | 7.97319 | 8.27176 | 7.70238 | 15.5114 | 20.8765 |
| 2011 | 8.37263 | 8.55315 | 8.19784 | 17.1275 | 22.554 |
| 2012 | 9.20598 | 9.42795 | 9.03972 | 18.9554 | 26.1072 |
| 2013 | 9.75897 | 10.1738 | 9.78391 | 21.2699 | 28.1667 |
| 2014 | 10.3364 | 10.6084 | 10.4061 | 22.086 | 30.2429 |
| 2015 | 10.1479 | 10.5105 | 10.4916 | 21.3767 | 29.274 |
| 2016 | 10.2965 | 10.8225 | 10.3288 | 22.2759 | 31.1139 |
| 2017 | 10.5305 | 10.9193 | 10.9098 | 22.9831 | 32.9117 |
| 2018 | 10.2678 | 10.8623 | 11.2816 | 23.97 | 33.7959 |
| 2019 | 10.141 | 10.7357 | 11.6924 | 23.7787 | 33.658 |

Figure S2b. Trend in CUD by Psychiatric Disorders, 35-64 Years

Patients with CUD Diagnosis, % (95% CI)

|  | Depressive Disorders | Anxiety Disorders | PTSD | Bipolar Disorders | Psychotic-Spectrum Disorders |
| --- | --- | --- | --- | --- | --- |
| 2005 | 4.25103 | 3.70842 | 3.98409 | 7.33665 | 6.06948 |
| 2006 | 4.30787 | 3.89867 | 3.89539 | 7.62904 | 6.50601 |
| 2007 | 4.40645 | 4.08757 | 4.00259 | 7.9775 | 6.68976 |
| 2008 | 4.65196 | 4.47999 | 4.11149 | 8.36979 | 7.06501 |
| 2009 | 5.09617 | 4.98611 | 4.51972 | 8.95033 | 7.9995 |
| 2010 | 5.50204 | 5.51686 | 4.94795 | 9.67669 | 8.78389 |
| 2011 | 5.75457 | 5.8677 | 5.24799 | 10.1879 | 9.51571 |
| 2012 | 6.10432 | 6.52559 | 5.80786 | 11.2557 | 10.8308 |
| 2013 | 6.48508 | 7.1352 | 6.35916 | 12.0837 | 11.8098 |
| 2014 | 6.9224 | 7.46886 | 6.82352 | 13.098 | 12.8936 |
| 2015 | 6.71071 | 7.29426 | 6.63332 | 12.9201 | 12.9774 |
| 2016 | 6.20564 | 7.00896 | 6.05265 | 12.6095 | 12.6068 |
| 2017 | 6.40263 | 7.26375 | 6.26384 | 13.637 | 13.5732 |
| 2018 | 6.44879 | 7.41823 | 6.39836 | 14.3321 | 14.5397 |
| 2019 | 6.57816 | 7.5627 | 6.61421 | 14.7672 | 15.693 |

Figure S2c. Trend in CUD by Psychiatric Disorders, 65+ Years

Patients with CUD Diagnosis, % (95% CI)

|  | Depressive Disorders | Anxiety Disorders | PTSD | Bipolar Disorders | Psychotic-Spectrum Disorders |
| --- | --- | --- | --- | --- | --- |
| 2005 | 0.22404 | 0.13948 | 0.38023 | 0.55584 | 0.35303 |
| 2006 | 0.27751 | 0.23657 | 0.41897 | 0.72206 | 0.45175 |
| 2007 | 0.36748 | 0.25732 | 0.51147 | 0.96796 | 0.66012 |
| 2008 | 0.47367 | 0.35597 | 0.5945 | 1.02941 | 0.73397 |
| 2009 | 0.61664 | 0.44826 | 0.7554 | 1.20825 | 0.93252 |
| 2010 | 0.76414 | 0.58074 | 0.98846 | 1.74639 | 1.16742 |
| 2011 | 0.91643 | 0.80695 | 1.12464 | 1.81985 | 1.42988 |
| 2012 | 1.12236 | 1.00018 | 1.36451 | 2.48091 | 1.91253 |
| 2013 | 1.40551 | 1.37008 | 1.70462 | 3.03537 | 2.45922 |
| 2014 | 1.71589 | 1.64091 | 2.08436 | 3.74097 | 2.82124 |
| 2015 | 1.90574 | 1.7724 | 2.31529 | 4.09388 | 2.95627 |
| 2016 | 1.98144 | 1.96458 | 2.21876 | 4.42367 | 3.10944 |
| 2017 | 2.33639 | 2.27878 | 2.65652 | 4.98879 | 3.63665 |
| 2018 | 2.64053 | 2.67001 | 3.23451 | 5.84016 | 4.342 |
| 2019 | 2.98902 | 3.1433 | 3.77408 | 6.71162 | 4.84618 |

^1^ Disorder categories are not mutually exclusive.

Hatch marks at 2015 indicate that this year was not included in models due to a change in ICD coding.
